# Polygenic risk score from a multi-ancestry GWAS uncovers susceptibility of heart failure

**DOI:** 10.1101/2021.12.06.21267389

**Authors:** Kuan-Han H. Wu, Nicholas J. Douville, Matthew C. Konerman, Michael R. Mathis, Scott L. Hummel, Brooke N. Wolford, Ida Surakka, Sarah E. Graham, Hyeon Joo, Jibril Hirbo, Nancy J. Cox, Simon Lee, Michael Preuss, Ruth J.F. Loos, Mark J. Daly, Benjamin M. Neale, Wei Zhou, Whitney E. Hornsby, Cristen. J. Willer, on behalf of the Global Biobank Meta-analysis Initiative

## Abstract

Identifying individuals at high risk of heart failure during precursor stages could allow for earlier initiation of treatments to modify disease progression. We performed a GWAS meta-analysis to generate a heart failure (HF) polygenic risk score (PRS) then tested the association with phenotypic subtypes (reduced ejection fraction [HFrEF] and preserved ejection fraction [HFpEF]) to evaluate the value of polygenic risk prediction. Results from the European-ancestry analysis showed that an ancestry-matched PRS, calculated from GBMI meta-analysis outperformed the previous HF GWAS (HERMES), yielding an adjusted odds ratio (aOR) of 2.27 (95% CI: 2.05-2.51; p: 1.76×10^−56^) from GBMI compared to 1.30 (95% CI: 1.18-1.44; p: 1.42×10^− 7^) from HERMES, and 1.49 (95% CI: 1.33-1.66; p: 8.38×10^−13^) compared to 1.17 (95% CI: 1.05- 1.31; p: 0.004) for HFrEF and HFpEF, respectively. Next, we evaluated the performance differences between ancestry-matched and multi-ancestry PRS in the African American cohort. The GBMI multi-ancestry GWAS-based PRS had a significant aOR of 1.49 (p: 0.006). Findings suggest that a PRS for heart failure derived from the GBMI multi-ancestry study is useful in predicting HFrEF, but less powerful in predicting HFpEF in an independent cohort. The difficulty in predicting HFpEF could result from the GBMI HF phenotype, preferencing HFrEF over HFpEF, and/or greater genetic heterogeneity in the HFpEF phenotype.

## INTRODUCTION

More than 26 million individuals globally are living with heart failure, which is a highly heterogeneous and progressive syndrome, resulting in the heart’s inability to deliver adequate blood flow to the body at normal filling pressures^1,2^. Heart failure is typically classified into phenotypic subtypes: (i) heart failure with a reduced ejection fraction (HFrEF) and (ii) heart failure with a preserved ejection fraction (HFpEF), based upon the left ventricular ejection fraction as a key distinction^3^. This classification provides a useful clinical distinction when diagnosing and managing patients with heart failure, given evidence-based therapies unique to each subtype.

Identifying individuals at a high risk of heart failure at early or precursor stages could allow for earlier initiation of treatments to modify disease progression^3^. Prior work suggests a genetic basis for heart failure secondary to varied etiologies, ranging from ischemic disease, hypertension, or cardiac arrhythmias^4^, but the genetics of heart failure is not fully understood. Utilization of large biobanks with genetic data, integrated with electronic health records, has the potential to identify large numbers of cases to improve statistical power, introduce greater genetic diversity, and balance varying etiologies.

To expand upon our current understanding of the genetics underpinning heart failure we calculated a polygenic risk score (PRS) derived from a new genome-wide association study (GWAS) for overall heart failure from the Global Biobank Meta-analysis Initiative (GBMI)^5^. GBMI is a global collaboration among 21 biobanks across the world with diverse ancestries. Next, we subtyped heart failure cases using a previously validated phenotyping algorithm^3^ to separately evaluate the association between the heart failure PRS and subtypes (HFrEF and HFpEF) in a large electronic health record-linked biobank. Findings from this study will elucidate the potential need for heart failure subtype-specific GWAS studies.

## RESULTS

The GBMI multi-ancestry heart failure meta-analysis marks the largest diverse heart failure genome-wide association study to date^6,7^. The meta-analysis included a total of 68,408 heart failure patients and 1,286,331 controls (5.1% cases) across six ancestral populations: 24.7% of the samples were of non-European-ancestry. Supplementary Table 1 describes the six ancestral populations included in the analysis. The prevalence of heart failure in our study cohorts ranged from 0.36% to 22.83%, with hospital-based biobanks contributing a larger number of cases (e.g., Mass General Brigham: 22.83%), compared to population-based cohorts (e.g., UKBB: 1.79% and HUNT: 0.36%), which are more representative of heart failure rates in the general population (0.3% to 2.1%, Supplementary Figure 1).

### GBMI Meta-Analysis Yields 14 Potentially Novel Loci for Heart Failure

Twenty-two independent loci reached genome-wide significance (p-value < 5×10-8) in the heart failure meta-analysis. Of the 22, 14 are putatively novel loci (Table 1) based on literature review and overlap with variants in the NHGRI-EBI GWAS Catalog^8^. Two of these loci, rs147288039 and rs373205748, were significant only in the multi-ancestry meta-analysis, likely due to higher allele frequency in East Asians (rs147288039: 0.23%) and South Asians (rs147288039: 0.75%; rs373205748: 0.08%) according to gnomAD.^9^ The inclusion of non-European ancestry samples has aided the genetic discovery for heart failure, demonstrating the power of genetic diversity and the importance of including multi-ancestry individuals to account for the genetic heterogeneity across populations.

**Table 1.** Variants significantly associated with heart failure outcome in GBMI. Twenty-two independent loci reached genome-wide significance, and of those, 11 are potentially novel loci.

| rsid | chr | pos (hg38) | ref | alt | nearest gene | function | beta | se | novel |
| --- | --- | --- | --- | --- | --- | --- | --- | --- | --- |
|  | 1 | 10736490 | G | A | CASZ1 | intronic | 0.037861 | 0.0066375 | 1 |
| rs74853338 | 2 | 200306928 | C | T | SPATS2L | intronic | 0.043398 | 0.0077942 | 0 |
|  | 3 | 27450659 | G | C | SLC4A7 | intronic | -0.035517 | 0.0064919 | 0 |
|  | 4 | 45173674 | C | T | GNPDA2;GABRG1 | intergenic | 0.040491 | 0.0067802 | 0 |
| rs201194999 | 4 | 65801177 | C | T | EPHA5-AS1;MIR1269A | intergenic | 0.11876 | 0.017742 | 0 |
| rs59788391 | 4 | 110780277 | A | G | PITX2;MIR297 | intergenic | 0.084207 | 0.0082364 | 1 |
| rs144757939 | 6 | 32638945 | A | G | HLA-DQA1 | intronic | 0.17803 | 0.028075 | 0 |
|  | 6 | 36665292 | A | G | MIR3925;PANDAR | intergenic | 0.053251 | 0.0066481 | 1 |
| rs10455872 | 6 | 160589086 | A | G | LPA | intronic | 0.11551 | 0.015208 | 1 |
| rs7857118 | 9 | 22124141 | A | T | CDKN2B-AS1;DMRTA1 | intergenic | 0.043202 | 0.0064814 | 1 |
| rs147288039 | 9 | 95006476 | A | G | AOPEP | intronic | 0.40002 | 0.070586 | 0 |
| rs600038 | 9 | 133276354 | C | T | ABO;SURF6 | intergenic | -0.051976 | 0.0074204 | 1 |
| rs373205748 | 10 | 103575604 | C | T | NEURL1 | intronic | 0.46961 | 0.081342 | 0 |
|  | 10 | 119665450 | A | T | BAG3 | intronic | -0.051487 | 0.0086631 | 1 |
| rs10774624 | 12 | 111395984 | G | A | PHETA1;SH2B3 | intergenic | -0.043648 | 0.0074662 | 1 |
|  | 12 | 131295306 | C | T | LINC02415 | downstream | 2.0587 | 0.35494 | 1 |
| rs62048402 | 16 | 53769311 | G | A | FTO | intronic | 0.050284 | 0.0065836 | 1 |
| rs61208973 | 16 | 72991194 | C | T | ZFHX3 | intronic | 0.045775 | 0.0073977 | 1 |
|  | 18 | 1821016 | A | T | LINC00470;METTL4 | intergenic | 0.066938 | 0.012151 | 0 |
| rs1788784 | 18 | 23579666 | A | G | NPC1 | intronic | -0.044129 | 0.0072417 | 0 |
| rs145478347 | 19 | 49671626 | G | A | BCL2L12 | intronic | 0.72167 | 0.11751 | 0 |
| rs558658474 | 20 | 49472840 | TC | T |  |  | -0.2957 | 0.050571 | 0 |

### GBMI Polygenic Risk Score

We next compared PRS generated from the present GBMI HF meta-analysis with the PRS generated from the previous HERMES GWAS (47,309 cases and 930,014 controls) to examine the change in PRS prediction with increasing GWAS sample size and evaluate the performance of genetic research utilizing large scale EHR-linked biobank^5^. The GBMI PRS outperformed the HERMES PRS, which is the largest publicly available heart failure GWAS to date. We restricted our validation cohort to European American individuals only in our Michigan Genomics Initiative (MGI)/ Cardiovascular Health Improvement Project (CHIP) combined cohort to compare the model performance of ancestry-matched PRS from GBMI (GBMI-EUR) and HERMES (HERMES-EUR) and multi-ancestry PRS from GBMI (GBMI-ALL). Both phenotyping driven HFrEF and HFpEF outcomes were significantly associated with all three PRSs in EA; furthermore, the ancestry-matched PRS built from GBMI meta-analysis performed best (Figure 1). To predict HFrEF, GBMI-EUR PRS yielded an adjusted odds ratio (aOR) of 2.27 (95% CI: 2.05-2.51; p-value: 1.76×10^−56^) per one standard deviation of normalized PRS increased, which was a significantly stronger predictor (non-overlapping confidence intervals) compared to HERMES-EUR PRS (aOR: 1.30 [95% CI: 1.18-1.44; p-value: 1.42×10^−7^]). Similar results were obtained in HFpEF: GBMI-EUR PRS had an aOR of 1.49 (95% CI: 1.33-1.66; p-value: 8.38×10^− 13^), compared to the HERMES-EUR PRS with an aOR of 1.17 (95% CI: 1.05-1.31; p-value: 0.004).

**Figure 1.**
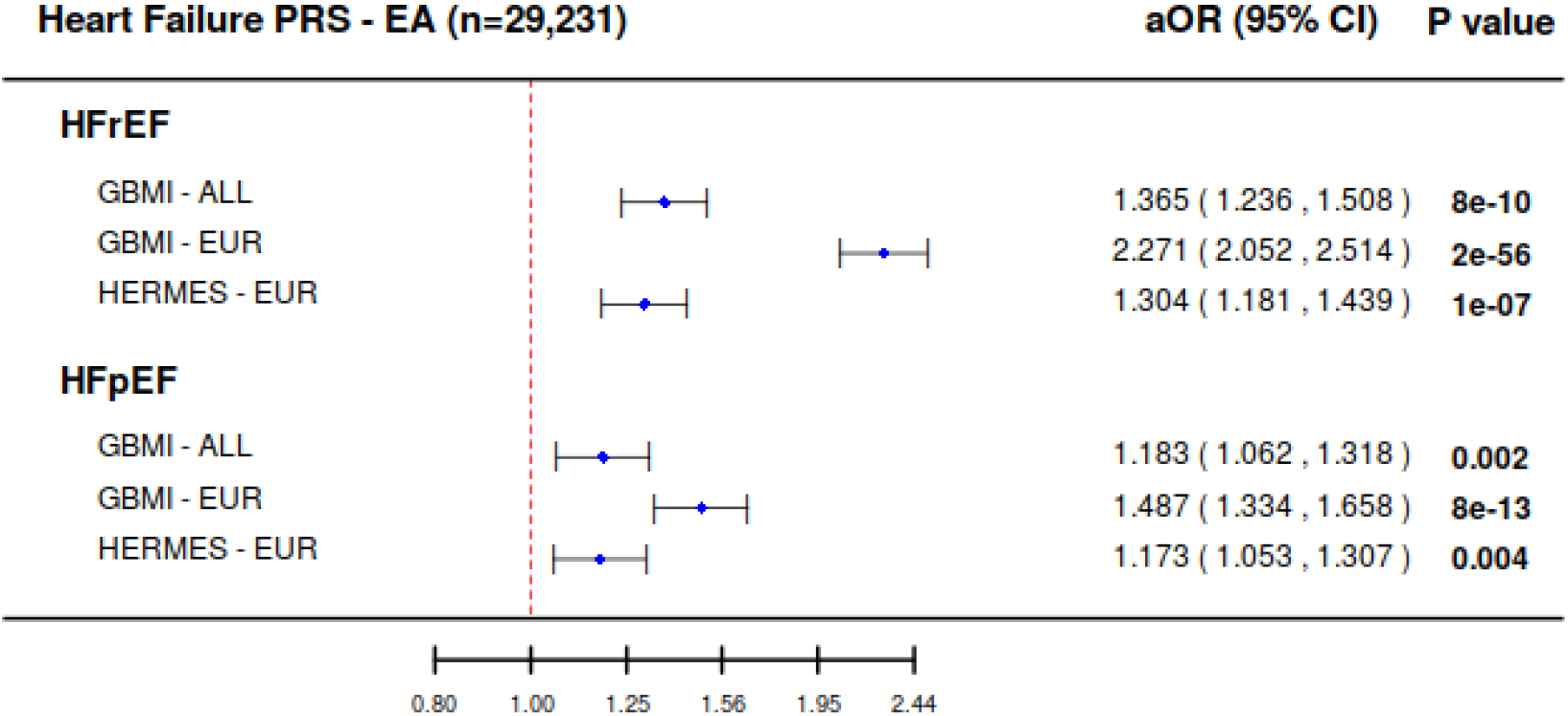
Forest plot of adjusted odds ratio comparison between heart failure PRS derived from GBMI-ALL, GBMI-EUR, and HERMES-EUR meta-analysis for HFrEF and HFpEF in European American. The GBMI PRS outperformed the HERMES PRS. Both HFrEF and HFpEF outcomes were significantly associated with heart failure PRS in European American; furthermore, ancestry-matched PRS built from GBMI meta-analysis performed optimally. GBMI-EUR PRS predicts cases of HFrEF, but notably less for cases of HFpEF.

Second, all PRSs better predicted cases of heart failure with reduced ejection fraction (HFrEF) than for preserved ejection fraction (HFpEF). In Figure 1, higher aORs were observed for HFrEF compared to HFpEF. Notably, the PRS derived from GBMI-EUR yielded a significantly stronger association with HFrEF (aOR: 2.26 [95% CI: 2.05-2.51]) compared to HFpEF (aOR: 1.49 [95% CI: 1.33-1.66]).

### The Effect of Genetic Diversity in Genome-wide Association Studies of Heart Failure

Given the determination that the GBMI PRS performed reasonably well in Americans with primarily European ancestry. We opted to further evaluate the ancestry transferability of PRS in the African American cohort, three separate PRS were created using the GBMI meta-analysis from different ancestral populations (1) multi-ancestry cohort (GBMI-ALL), (2) European ancestry-only cohort (GBMI-EUR), and (3) African ancestry-only cohort (GBMI-AFR) GWAS meta-analyses. We observed that the multi-ancestry score improved the model performance in the AA cohort (Figure 2). The same trend of ancestry-matched PRS yielding the best performance in the EA cohort was not observed in the AA cohort, possibly due to smaller sample size in GBMI-AFR GWAS (n=31,202) (Supplementary Figure 2; Supplementary Table 1 & 2). The best performing PRS in the AA cohort was the multi-ancestry score, which had a significant aOR of 1.49 (95% CI: 1.12-1.98; p-value=0.006) in HFrEF and the highest, although nonsignificant aOR of 1.33 (95% CI: 0.94-1.87; p-value: 0.11) in HFpEF. Neither the ancestry-matched score (GBMI-AFR) nor the EA-best performing score (GBMI-EUR) were significantly associated with heart failure outcome in AA. These findings demonstrate that the trans-ancestry based PRS is useful in predicting HFrEF in both EA and AA cohorts. Consistently, a less predictive trend using general HF PRS to predict HF subtype was observed for HFpEF in the African American cohort, as well.

**Figure 2.**
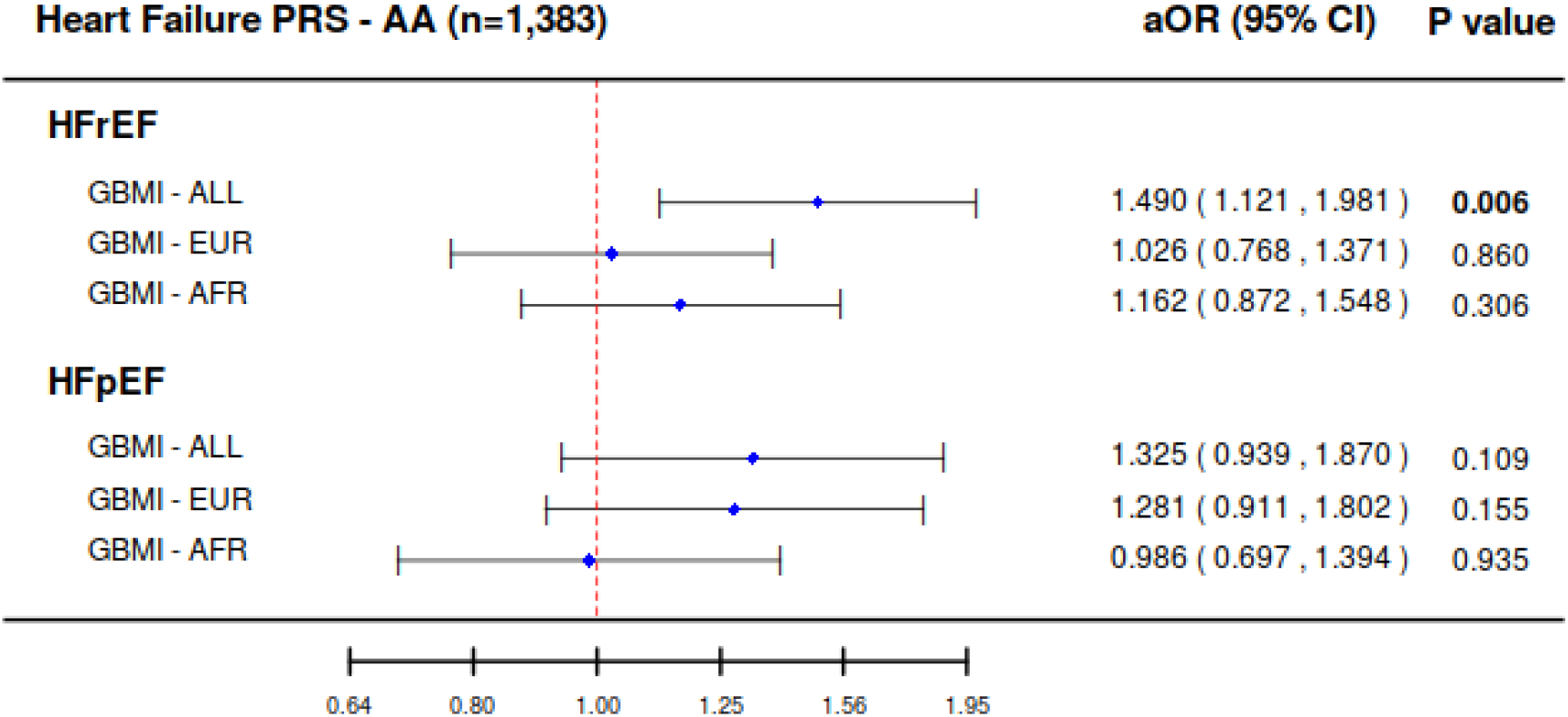
Forest plot of adjusted odds ratio comparison between heart failure PRS derived from GBMI-ALL, GBMI-EUR, and HERMES-AFR meta-analysis for HFrEF and HFpEF in African American. Multi-ancestry score improved the model performance in the African American cohort, compared among (1) multi-ancestry cohort (GBMI-ALL), (2) European ancestry-only cohort (GBMI-EUR), and (3) African ancestry-only cohort (GBMI-AFR) meta-analysis GWAS results.

### Pleiotropic Effect of Heart Failure Genetic Variants

Phenome wide association study (PheWAS) in UK Biobank white British cohort revealed association between the heart failure PRS and other cardiovascular diseases. The result showed that the heart failure PRS was associated with increased risk of: hypertension (aOR=1.05; p=1.72×10^−26^), coronary atherosclerosis (aOR=1.05; p=1.33×10^−9^), and atrial fibrillation (aOR=1.04; p=6.89×10^−7^). Additionally, the PheWAS demonstrated pleiotropy between the PRS for heart failure and increased risk of complex, systemic disease processes including obesity (aOR=1.04; p=5.36×10^−6^) and diabetes mellitus (aOR=1.03; p=7.97×10^−6^).

## DISCUSSION

Genome-wide discovery for heart failure traits based on 68,408 cases and 1,286,331 controls from six ancestry groups identified 22 index variants (14 novel) reaching genome-wide significance. A high proportion of the 22 index variants identified in our study were previously reported in GWAS Catalog to be associated with cardiovascular diseases. We further investigated the effect of genetically-predicted heart failure outcome on other diseases and conditions in the PheWAS, and confirmed known pleiotropic associations were confirmed with other cardiovascular phenotypes, such as hypertension, atrial fibrillation, and coronary atherosclerosis (Figure 3). These likely occur through a combination of both *biological*-pleiotropy (the genetic underpinning influences more than one phenotype) and *mediated*-pleiotropy (the phenotype itself is causally related to a second phenotype).^10^

**Figure 3.**
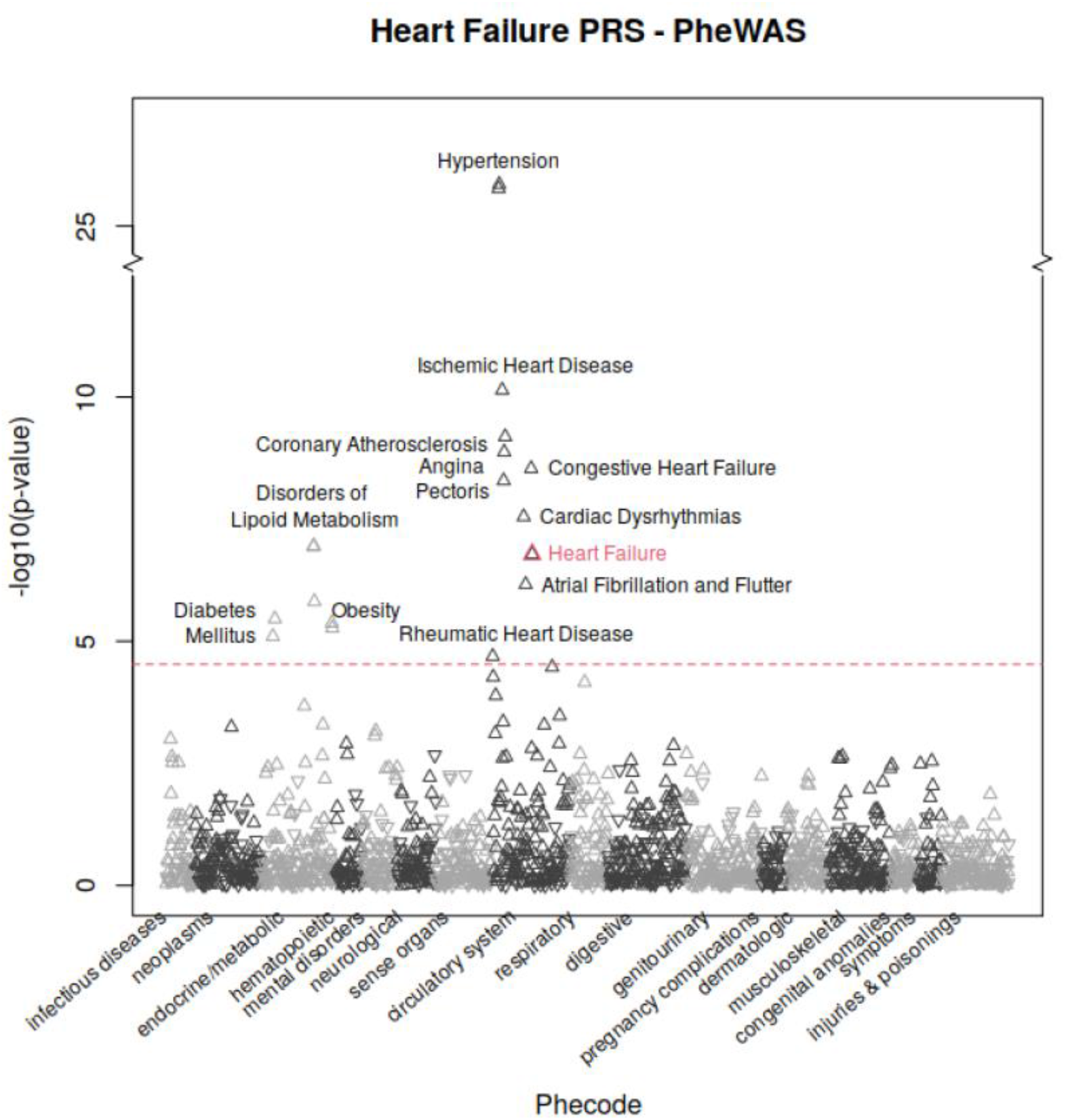
Manhattan plot of heart failure PRS PheWAS presenting the association between heart failure PRS and 1,688 phecode. Phenome-wide association study in the UK Biobank white British cohort revealed pleiotropic clustering between the heart failure PRS and other cardiovascular diseases. Positive associations were indicated by upward pointing triangles and negative associations were indicated by downward pointing triangles. Phecode 428.2 (heart failure), primary outcome of this study, was highlighted in red. Significance level was indicated by red dotted line with bonferroni correction for 1,688 tests, at 4.53, -log10(0.05/1688).

Heart failure may result from varied etiology, including: (i) ischemic disease, (ii) valvular abnormalities (as can be caused by rheumatic heart disease, bicuspid aortic valve, senile calcific stenosis, or endocarditis), (iii) arrhythmias, and (iv) hypertension.^11^ Therefore, the pleiotropic clustering with each of these diseases likely *mediating* heart failure is also not surprising. Additionally, identified phenotypes may themselves be both precipitating and secondary processes, as with the pathophysiologic cycle between atrial fibrillation and heart failure (a complex association, explored in depth by Anter, Jessup, and Callans).^12^ The link between diabetes mellitus, obesity, and disorders of lipid metabolism with heart failure likely results from *biological*-pleiotropy and is more complex than a sample causal association.^13^ Evidence from genetic epidemiology suggest genomic loci exert pleiotropic effects on multiple cardiovascular risk factors, including: (i) diabetes mellitus,^14–16^ (ii) obesity,^17,18^ and (iii) dyslipidemia.^19,20^ Therefore, the clustering demonstrated in our PheWAS between heart failure and a variety of cardiovascular diseases (and risk factors for cardiovascular diseases) is expected and explainable through a overlapping mechanisms.

Next, in comparison with a PRS constructed from the previous largest heart failure GWAS -- HERMES -- we show that increased GWAS sample sizes generate a heart failure PRS that can better identify heart failure cases of both subtypes, HFrEF and HFpEF, highlighting the additive power of large sample sizes. The GBMI PRS outperformed HERMES at identifying heart failure cases of both subtypes, although we observed enhanced diagnostic accuracy for HFrEF compared to HFpEF within our validation dataset. This could be due to a combination of factors, including: (i) the GBMI heart failure phenotype capturing HFrEF over HFpEF, (ii) greater statistical power to test the PRS due to a larger number of cases in the HFrEF cohort, and (iii) a stronger genetic association with HFrEF versus HFpEF (i.e., greater genetic heterogeneity in the HFpEF population). First, to test our hypothesis for phenotype preferencing, we sub-categorized heart failure subtypes within BioVU and BioMe (GBMI study cohorts) using phecode 428.3 for HFrEF and 428.4 for HFpEF. We found by using ICD code classification in two discovery cohort that approximately 58% of the heart failure cases were HFrEF patients. Similarly, we observed a higher proportion of HFrEF cases (55%) within our MGI/CHIP combined cohort. Second, our findings that PRS predicts HFrEF may be due to a large number of cases in MGI/CHIP (453 cases of HFrEF versus 369 cases of HFpEF) and larger sample size in the European American cohort (29,231 EA versus 1,383 AA individuals), leading to greater statistical power to distinguish between those with a high or low genetic risk. However, given the approximate similarity in case numbers with HFrEF and HFpEF in the discovery and testing cohorts, we expect that these issues may not be the factors driving the substantial difference in PRS performance by heart failure subtypes.

Third, studies have shown that disease subtypes could potentially have distinct genetic risk^21,22^ or different effect sizes among disease sub-category. According to Pividori et al.,^21^ genetic variants identified from an adult-onset asthma GWAS overlap with loci identified from childhood-onset asthma GWAS, but the effect sizes were significantly different by asthma endotypes.^21^ They observed larger genetic effects related to childhood-onset asthma, suggesting that genetic risk plays an important role in childhood-onset asthma, whereas environmental risk contributes to adult-onset asthma. Disease endotypes having distinct genetic architecture were also reported for polycystic ovary syndrome by Dapas et al.^22^ GWAS findings show independent loci associated with reproductive (4 loci) and metabolic (1 locus) polycystic ovary syndrome subtypes, respectively.^22^ These studies highlight the importance of using phenotypic subtyping to understand genetic nuances underlying various diseases. Thus, we postulate that there may be a stronger genetic association with HFrEF versus HFpEF (or greater genetic heterogeneity in the HFpEF population).

HFpEF is a heterogeneous disease with multiple different phenotypes.^23,24^ First, several comorbid conditions such as hypertension, diabetes mellitus, obesity, and others have been implicated in the pathophysiologic mechanisms driving HFpEF development and progression.^25– 27^ Patients with HFpEF can have some but not all of these comorbid conditions. These conditions each may have their own genotypic characteristics that could make isolating any one HFpEF genotype more difficult. Second, numerous pathophysiologic mechanisms have been implicated in the disease involving abnormalities in the left ventricular myocardium, left atrium, pulmonary vasculature, arterial stiffness, and skeletal muscle.^26,28–33^ Lastly, the diagnostic criteria used in guidelines and clinical trials have varied.^34^ Patients can have HFpEF despite not meeting all diagnostic criteria for the disease.^35,36^ For example, patients with obesity may have HFpEF without elevated natriuretic peptide levels.^37–39^ Unlike HFrEF, the diagnosis of HFpEF cannot rely on a reduced ejection fraction as a defining characteristic of the disease. For all of the above reasons, many have argued that treatments for this heterogeneous disease must be targeted to specific phenotypes.^23–25^ Thus, it is reasonable to conclude that specific HFpEF phenotypes may have specific genetic causes. However, identifying these genotypes requires a granular classification of HFpEF phenotypes not easily achieved in retrospective analyses of large datasets. Taken together, any or a combination of these factors may have contributed to the PRS in our study being less powerful in predicting HFpEF.

### Limitations

Beyond the limitations noted above, this study is also limited by reduced sample size in the GBMI African ancestry meta-analysis (supplementary figure 2) and reduced sample size in the MGI/CHIP combined cohort. Moreover, we do not have enough individuals with East Asian, South Asian, or Admixed/ Latino American ancestry in our dataset to validate PRS transferability in different ancestral cohorts. The low performance of ancestry-matched PRS score in AA (AFR meta-analysis [1,230 cases; 27,092 controls]; AFR individuals in MGI/CHIP [50 HFrEF; 34 HFpEF]) could potentially be due to lower discovery GWAS sample size, compared to EA ancestry (EUR meta-analysis [51,274 cases; 922,900 controls]; EUR individuals in MGI/CHIP [403 HFrEF; 335 HFpEF]). Studies with comparable sample sizes in both training and testing sets are needed to examine the effect from ancestry-match and multi-ancestry PRS.

Also, the low performance in HFpEF for AA potentially could be due to lower proportion of HFpEF in AA. We observed that AA have a higher proportion of HFrEF (59% HFrEF) compared to EA (54% HFrEF). All three biobanks (MGI/CHIP, BioVU, and BioME) consistently contributed a higher proportion of HFrEF cases in AA than EA.

### Outlook/Conclusion

This study investigated genetic-based prediction of heart failure within subtypes and the power of sample size and diverse ancestry in GWAS. In the future, generating higher quality phenotypes (perhaps more defined than just ICD-9/10 codes) could further unravel the genetic underpinnings of subtype-specific genetic risks, particularly in the case of heart failure where the two subtypes show substantial differences in performance of genetic prediction tools. Secondly, GWAS with larger sample sizes could likely increase the loci discovered and improve our understanding of the biology at established loci. Together, these approaches may more efficiently identify traits in early or precursor stages, allowing for early initiation of treatments to augment disease progression.

## Data Availability

Meta-analysis conducted by Global Biobank Meta-analysis Initiative are available online at http://results.globalbiobankmeta.org/

## Acknowledgments

We would like to express our gratitude to all contributors to GBMI and the biobank participants who provided their data for biomedical research. Particularly, we are grateful to the GBMI study cohorts BioVU and BioME for further sub-categorizing heart failure subtypes to assist with the interpretation of these findings. The authors acknowledge the participants, recruitment teams and project managers of the Global Biobank Meta-analysis Initiative for providing data aggregation, management, and distribution services in support of the research reported in this publication (particularly Sinéad Chapman and Bethany Klunder).

We acknowledge study team members from the participating cohorts: BioBank Japan (Yukinori Okada, Koichi Matsua, and Masahiro Kanai), BioMe (Ruth Loos, Judy Cho, Eimear Kenny, Michael Preuss, and Simon Lee), BioVU (Nancy Cox, Jibril Hirbo, and Megan Shuey), Canadian Partnership for Tomorrow (Philip Awadalla and Marie-Julie Fave), China Kadoorie (Robin Walters, Kuang Lin, and Iona Millwood), Colorado Center for Personalized Medicine (Kathleen Barnes, Michelle Daya, and Chris Gignoux), deCODE Genetics (Kári Stefánsson and Unnur Þorsteinsdóttir), East London Genes & Health (David A van Heel, Sarah Finer, and Richard Trembath), Estonian Biobank (Andres Metspalu, Reedik Mägi, Tõnu Esko, and Priit Palta), FinnGen (Aarno Palotie, Mark Daly, Samuli Ripatti, Mitja Kurki, and Juha Karjalainen), Generation Scotland (Caroline Hayward and Riccardo Marioni), HUNT (Kristian Hveem, Cristen Willer, and Sarah Graham, Ben Brumpton, and Brooke Wolford), Lifelines (Serena Sanna and Esteban Lopera), Michigan Genomics Initiative (Sebastian Zoellner, Michael Boehnke, Lars Fritsche, and Anita Pandit), Million Veteran Program (Christopher J. O’Donnell), Netherlands Twin Register (Dl Boomsma, MG Nivard), Partners Biobank (Jordan Smoller and Yen-Chen Feng), QIMR Berghofer (Sarah Medland, Stuart McGregor, and Nathan Ingold), Taiwan Biobank (Yen-Feng Lin, Yen-Chen Feng, and Hailiang Huang), UCLA Precision Health Biobank (Ruth Johnson, Yi Ding, Alec Chiu, Bogdan Pasaniuc, and Daniel Geschwind), and UK Biobank (Konrad Karczewski and Alicia Martin).

## Author Contributions

Bioinformatic and statistical analysis: WZ, KHW, IS, BNW

Drafting the manuscript: KHW, WEH, NJD, MCK

Senior writing team: CJW, WEH

Critical revision of the manuscript: KHW, NJD, MCK, MRM, SLH, BNW, IS, SEG, HJ, JH, NJC, SL, MP, RJFL, MJD, BMN, WZ, WEH, CJW

## Declaration of interests

C.J.W’s spouse works at Regeneron pharmaceuticals.

M.J.D. is a founder of Maze Therapeutics.

B.M.N. is a member of the scientific advisory board at Deep Genomics and consultant for

Camp4 Therapeutics, Takeda Pharmaceutical, and Biogen.

## STAR Methods

### Multi-Ancestry Meta-Analysis

Global Biobank Meta-analysis Initiative (GBMI) is a global collaboration among 21 biobanks across the world that aims to equitably impact people of diverse ancestries. Biobanks in GBMI reach across 4 continents and have more than 2.6 million individuals with electronic health record linked genetic information. Biobanks that contributed to heart failure study include BioBank Japan, BioMe, BioVU, China Kadoorie Biobank, Estonian Biobank, FinnGen, Genes & Health, HUNT, Lifelines, Michigan Genomics Initiative, Partners Biobank, UCLA Precision Health Biobank, and UK Biobank (Supplementary Figure 1). Heart failure cases in the GBMI training dataset were defined based upon ICD codes (phecode 428.2: heart failure, not otherwise specified), which did not distinguish between heart failure subtypes (Reference to GBMI flagship).^5^ In the GBMI discovery dataset, genetic data was analyzed from a total of 67,049 HF patients from 1,305,592 samples from 6 ancestral populations: 25.4% of the samples were of non-European ancestry (Supplementary Figure 1; Supplementary Table 1).

### Polygenic Risk Score

We aimed to compare the prediction accuracy of PRS derived from GBMI heart failure GWAS and the largest published heart failure GWAS conducted by Heart Failure Molecular Epidemiology for Therapeutic Targets (HERMES) Consortium.^6^ HERMES comprises 977,323 individuals of European ancestries, and of those, 4.8% are heart failure cases. Three PRSs were generated to compare the performance between GBMI and HERMES in European American: (i) GBMI with multi-ancestries cohort (GBMI - ALL; n=1,354739 [5.0% cases]), (ii) GBMI with European ancestry cohort (GBMI - EUR; n=1,020,441 [5.1% cases]), and (iii) HERMES with European ancestry cohort (HERMES - EUR; n=977,323 [4.8%]) (Supplementary Figure 2; Supplementary Table 1).

Additional analysis on PRS transferability was performed in the AA subset of MGI/CHIP to compare the model performance between PRS built from ancestry-specific and trans-ancestry meta-analysis. Three sets of PRS were derived from (i) GBMI multi-ancestry, (ii) GBMI European-ancestry, and (iii) GBMI African-ancestry (GBMI - AFR; n=31,202 [4.4% cases]) meta-analysis (Supplementary Figure 2; Supplementary Table 1).

Polygenic risk score weights were calculated using PRS-CS^40^ with a reference panel from the combined cohort of 1000 Genomes and UK Biobank. For multi-ancestry and European-ancestry GWAS, a LD panel from individuals of European-ancestry was used. For African-ancestry GWAS, a LD panel from the African-ancestry cohort was used. The summary statistics used to generate PRS weights in our main analysis excluded our testing cohort, MGI, and in phenome-wide association study to evaluate the pleiotropic effect of HF genetic risk excluded UK Biobank (Supplementary Table 2). To control for possible population structure, each of the six raw PRSs were further regressed on the top 10 principal components (PC) derived from the genotype data within each ancestry group. The resulting residuals were further transformed to normal distribution using inverse normalization within each ancestry group to generate the final heart failure PRSs for each individual..

### Statistical Analysis

Model performance for prediction accuracy of PRS derived from GBMI heart failure GWAS to one derived using the previously largest published summary statistics from HERMES were evaluated using European ancestry samples in MGI/CHIP. PRS ancestral transferability was tested in AA subset of MGI/CHIP by comparing the model performance between trans-ancestry, ancestry-matched, and ancestry-mismatched PRSs.

We evaluated logistic regression models with PRS adjusted for age and sex separately for both HFrEF and HFpEF phenotypes to compare the odds ratio of different PRSs. The significance levels were adjusted for multiple tests correction using Bonferroni adjustment. The corrected p-value threshold 0.008 (0.05/6) was corrected for the number of outcomes (2 subtypes; HFrEF and HFpEF) and the number of GWAS summary statistics (2 GWAS; GBMI and HERMES) and the number of ancestry-specific cohort (2 cohorts; European American and African American).

### Phenome-Wide Association Study (PheWAS)

Phenome-wide association study was conducted in 408,155 white British individuals from United Kingdom Biobank (UKBB).^41^ Logistic regression was performed to examine the association between disease status for 1,688 phecodes^42^ as dependent variable and heart failure PRS as independent variable. Models were adjusted for sex, birth year, and top four PCs derived from genotype file of the participants. Heart failure PRS calculated in UK Biobank was derived from leave UK Biobank cohort out meta-analysis from GBMI. Bonferroni correction was applied to account for multiple tests in PheWAS. Significance level was set to 2.96*10-5 for adjusting 1,688 tests (0.05/1688) in total.

Combined Cohort

Michigan Genomics Initiative (MGI), a longitudinal biorepository within Michigan Medicine, from 2014 to 2020. MGI has integrated genetic data with electronic health records (EHR) on adult patients (≥18 years) undergoing surgery within Michigan Medicine. The Cardiovascular Health Improvement Project (CHIP) Biorepository is a longitudinal observational cohort study of patients at Michigan Medicine, from 2013 to 2021, with a clinical diagnosis of cardiovascular disease (predominantly, thoracic/abdominal aortic disease or HFpEF).^43^ The University of Michigan’s Institutional Review Board approved these protocols (HUM00128472 and HUM00052866) and all study participants signed informed consent.

Individuals in the combined cohort with both electronic health records and genetic information available were included in our study. Patients with age or sex missing data were excluded. GWAS summary statistics were used to generate PRSs from the Michigan Genomics Initiative and Cardiovascular Health Improvement Project (MGI/CHIP). A total of 27,848 EA individuals (403 HFrEF cases and 335 HFpEF cases) were included in the primary analysis to compare the model performance among PRS built from GBMI-ALL, GBMI-EUR, and HERMES- EUR. For PRS transferability analysis, 1,383 AA samples (50 HFrEF and 34 HFpEF) were included. (Supplementary Table 3)

### Subtype Definition

We integrated two sources of label curation from MGI and CHIP to define a total of 453 HFrEF cases and 369 HFpEF cases. Electronic health record data enabled further classification of the patients into HFrEF, HFpEF, and healthy controls; using the previously validated methodology.^3^ In MGI, we used the previously published phenotyping algorithm^3^ and defined 453 and 279 patients with HFrEF and HFpEF, respectively. In CHIP, 90 HFpEF patients were assigned with a gold-standard label by manual label curation from HFpEF specialists (S.L.H. and M.C.K.).

The inclusion criteria for methodology applied in MGI for heart failure subtype definition was adult patients, ≥ 40 years of age, who had at least 2 episodes of care at Michigan Medicine from 2010 to 2019, and were enrolled within MGI. In brief, patients with a qualifying heart failure ICD-9/10 code (or code for cardiomyopathy or cardiomegaly) *and* LVEF ≤ 40% on cardiac imaging were classified as HFrEF. Patients with (i) a qualifying HF diagnostic code,* (ii) all LVEF > 50% (at least one LVEF available), and (iii) positive mention of heart failure keyword** within the EHR were classified as HFpEF. Patients with (i) no qualifying ICD-9/10 codes,*^1^ (ii) LVEF ≥ 50% on all available cardiac imaging (no requirement for LVEF study), (iii) no mention of heart failure keywords in EHR, and (iiv) not on any uniquely heart failure medications *** were classified as healthy controls. Data quality and heart failure subtype veracity were confirmed with adjudication by expert clinician review (N.J.D. and M.R.M.).^3^

The diagnosis of HFpEF in CHIP was made by cardiologists, sub specializing in HFpEF, based on the the 2016 European Society of Cardiology guidelines: i) Signs and/or symptoms of heart failure, ii) left ventricular ejection fraction ≥50%, at least mild elevation in natriuretic peptide levels, and iii) cardiac structural (e.g. left atrial enlargement) and/or functional abnormalities (e.g. diastolic dysfunction) associated with HFpEF^46^. Participants may have been diagnosed with HFpEF following hospitalization for decompensated HF requiring intravenous diuresis and/or if increased left ventricular filling pressures were documented on catheterization, regardless of natriuretic peptide level.

## Supplementary Figure

**Supplementary Figure 1.**
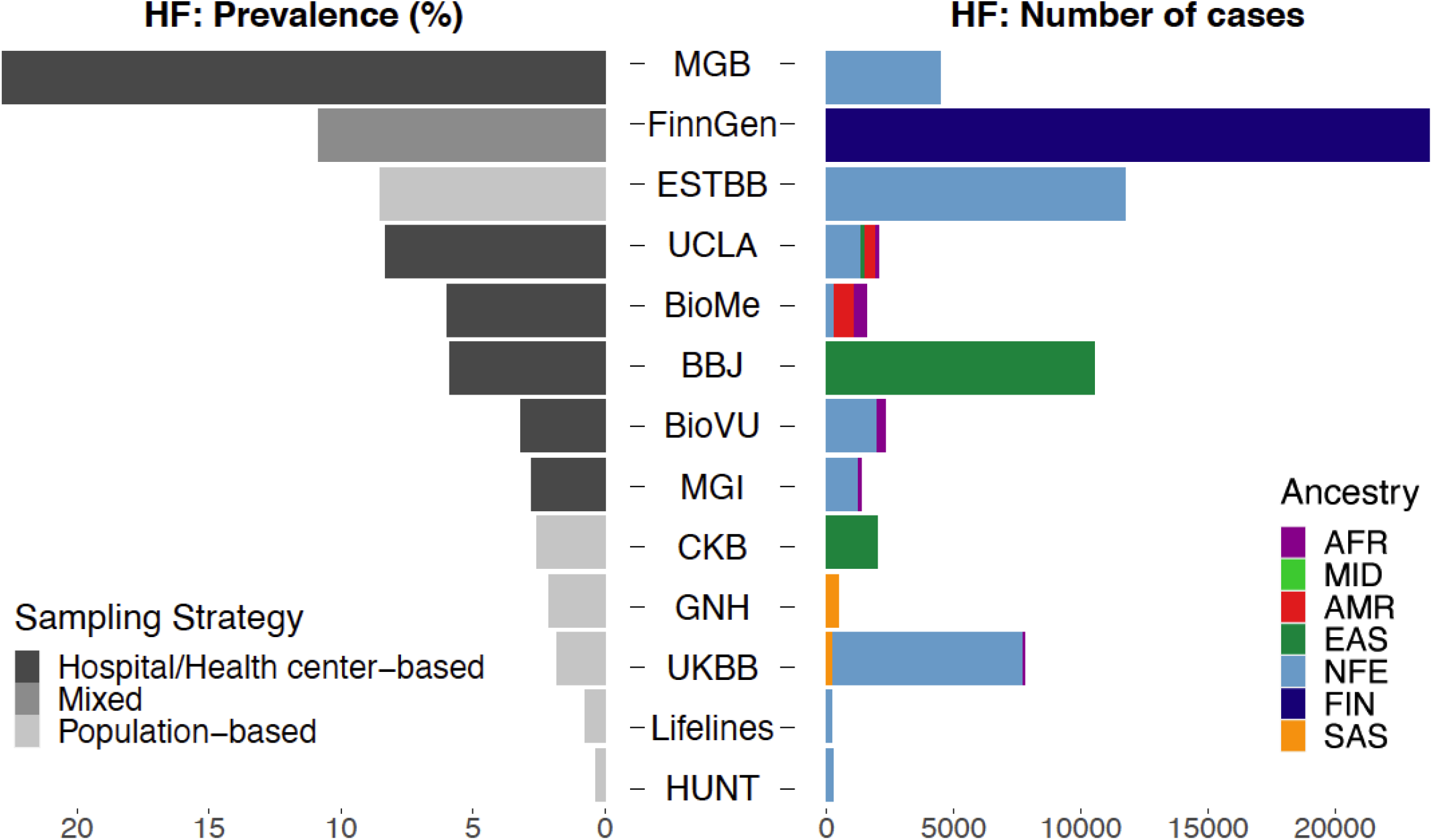
Sample sizes and heart failure prevalence across studies and ancestries. Left panel: prevalence of heart failure by biobank, recruitment strategies were indicated by the colors. Right panel: sample size within each ancestry by biobank. Biobanks were sorted by heart failure prevalence.

**Supplementary Figure 2.**
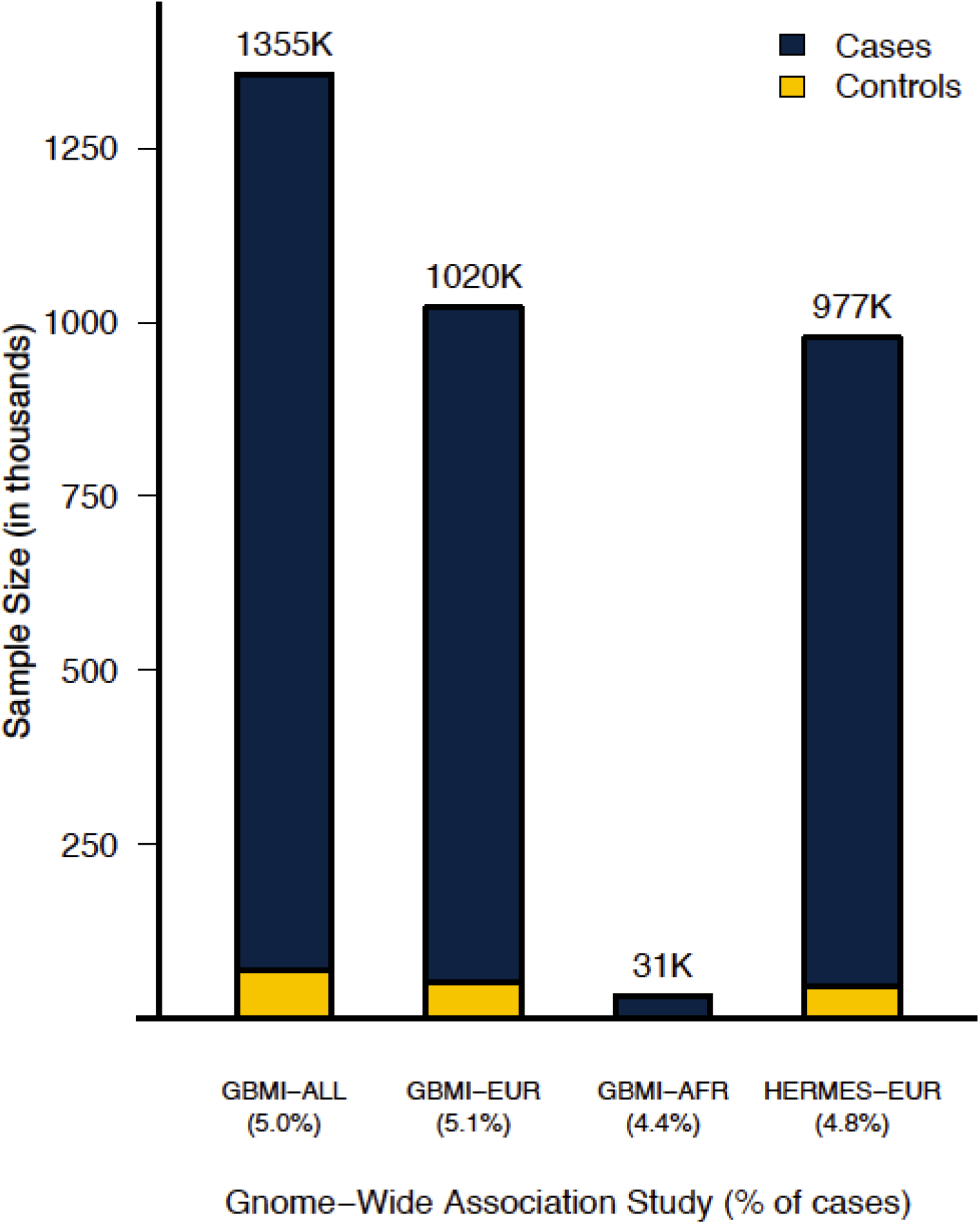
Barplot of total GWAS sample sizes and proportion of heart failure cases, total numbers of individuals in GWAS were indicated on the top of the bar. Comparison between sample size for GBMI (i) multi-ancestry, (ii) European ancestry, (iii) African ancestry, and HERMES (iv) European ancestry meta-analysis.

**Supplementary table 1.** Sample size across ancestries in all biobanks contributed to heart failure GWAS.

| Ancestries | GBMI* |  |  |
| --- | --- | --- | --- |
|  | Cases<sup>1</sup> | Controls<sup>1</sup> | Total<sup>2</sup> |
| African (AFR) | 1,367 (4.4%) | 29,835 (95.6%) | 31,202 (2.3%) |
| American (AMR) | 1,179 (8.1%) | 13,217 (91.9%) | 14,387 (1.1%) |
| East Asian (EAS) | 12,665 (4.9%) | 245,263 (95.1%) | 257,928 (19.0%) |
| Finnish (FIN) | 23,701 (10.8%) | 195,091 (89.2%) | 218,792 (16.1)) |
| Non-Finnish European (NFE) | 28,795 (3.6%) | 772,854 (94.4%) | 801,649 (59.2%) |
| South Asian (SAS) | 710 (2.3%) | 30,071 (97.7%) | 30,781 (2.3%) |
| Total | 68,408 (5.1%) | 1,286,331 (94.9%) | 1,354,739 |

\* GBMI cohort contributed to HF: BioBank Japan, BioMe, BioVU, China Kadoorie Biobank, Estonian Biobank, FinnGen, Genes & Health, HUNT, Lifelines, Michigan Genomics Initiative, Partners Biobank, UCLA Precision Health Biobank, and UK Biobank.
1 Percentage by total number of samples within each ancestry
2 Percentage by total number of individuals across all ancestries

**Supplementary Table 2.**
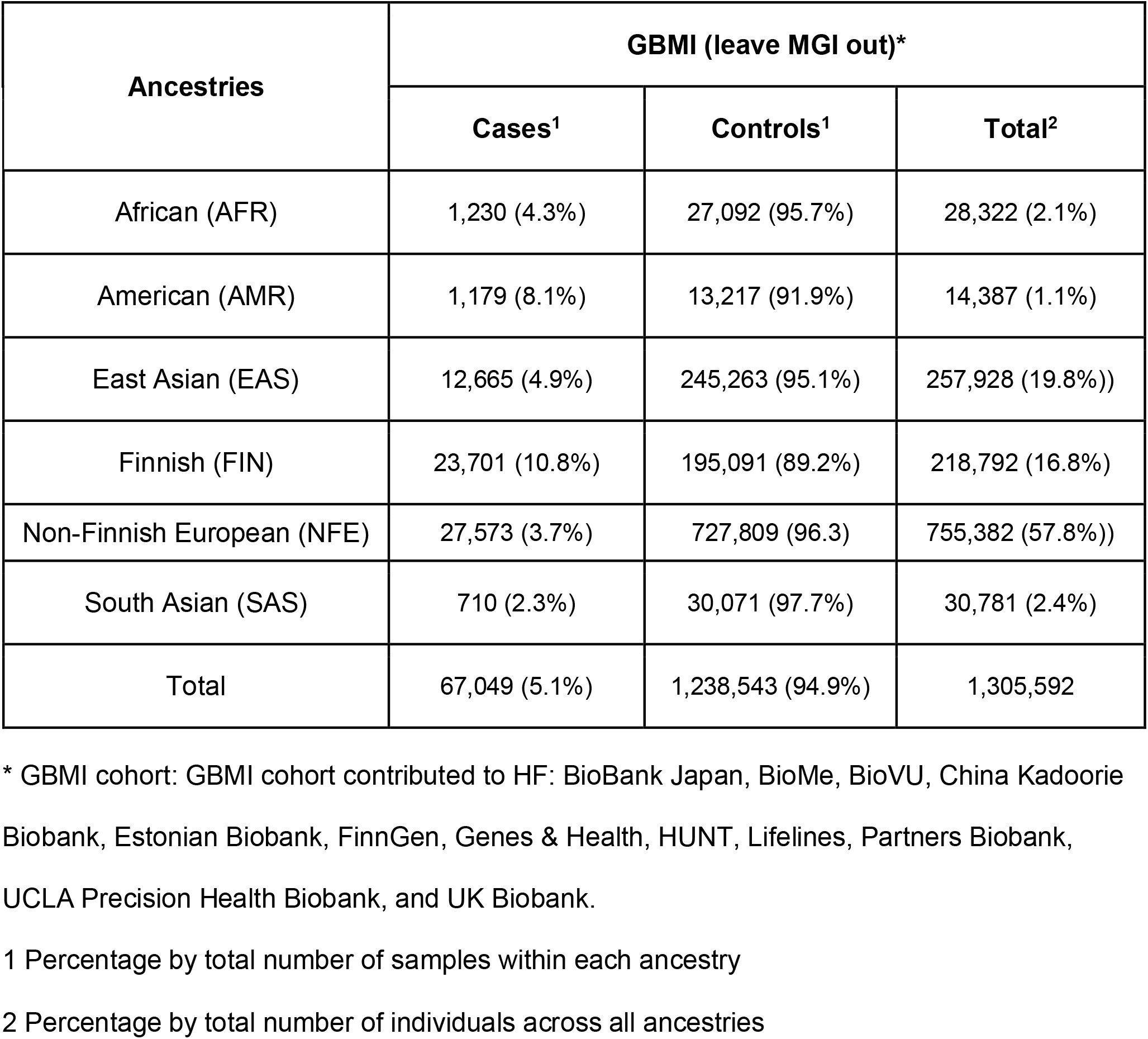
Sample size across ancestries in all biobanks, but MGI, contributed to heart failure GWAS.

| **Ancestries** | **GBMI (leave MGI out)\*** |  |  |
| --- | --- | --- | --- |
|  | **Cases<sup>1</sup>** | **Controls<sup>1</sup>** | **Total<sup>2</sup>** |
| African (AFR) | 1,230 (4.3%) | 27,092 (95.7%) | 28,322 (2.1%) |
| American (AMR) | 1,179 (8.1%) | 13,217 (91.9%) | 14,387 (1.1%) |
| East Asian (EAS) | 12,665 (4.9%) | 245,263 (95.1%) | 257,928 (19.8%)) |
| Finnish (FIN) | 23,701 (10.8%) | 195,091 (89.2%) | 218,792 (16.8%) |
| Non-Finnish European (NFE) | 27,573 (3.7%) | 727,809 (96.3) | 755,382 (57.8%)) |
| South Asian (SAS) | 710 (2.3%) | 30,071 (97.7%) | 30,781 (2.4%) |
| Total | 67,049 (5.1%) | 1,238,543 (94.9%) | 1,305,592 |

\* GBMI cohort: GBMI cohort contributed to HF: BioBank Japan, BioMe, BioVU, China Kadoorie Biobank, Estonian Biobank, FinnGen, Genes & Health, HUNT, Lifelines, Partners Biobank, UCLA Precision Health Biobank, and UK Biobank.
1 Percentage by total number of samples within each ancestry
2 Percentage by total number of individuals across all ancestries

**Supplementary Table 3.** Sample size by heart failure subtypes and demographic characteristics in MGI/ CHIP cohort.

|  | **Overall (n=29,231)** | **AA (n=1,383)** | **EA (n=27,848)** |
| --- | --- | --- | --- |
| HFrEF | 453 | 50 | 403 |
| HFpEF | 369 | 34 | 335 |
| Age | 60.27 ± 11.08 | 56.69 ± 10.21 | 60.44 ± 11.09 |
| Female | 14,995 | 833 | 14,162 |
| Male | 14,236 | 550 | 13,686 |

## Footnotes

1* Defined by Elixhauser Enhanced ICD-9-CM and ICD-10 Indices: Congestive Heart Failure (428.X (ICD-9) or I50.X (ICD-10), X *= wildcard*), as well as ICD-9 and ICD-10 codes for cardiomyopathy (425.X, I42.X) and cardiomegaly (429.3, I51.7)

** Heart Failure Keywords: “HF”, “heart fail*”, “cardiac fail*”, “ventricular fail*”, “cardiomyop*”, or “*ICM”; *\* = wildcard*

*** Uniquely Heart Failure Medications: Digoxin (Lanoxin), sacubitril/valsartan (Entresto)

